# Predicting the stage of gastric cancer after gastrectomy based on machine learning algorithms

**DOI:** 10.1101/2025.04.04.25325291

**Authors:** Nayereh Abdali, Sajad Alavimanesh, Mirhamid Mirsaeid Ghazi, Seyedeh Negin Hadisadegh

## Abstract

**Background:** Gastric cancer (GC) is the fourth most common cause of cancer death worldwide, with a 5-year survival rate of less than 40%. One of the most important methods for diagnosing stomach cancer is endoscopy, which is quite costly and invasive. The aim of this study was to develop machine learning-based diagnostic prediction models for the stage of GC.

**Objectives:** To create a highly accurate predictive model for the stage of GC in patients via a noninvasive method based on machine learning (ML).

**Methods:** In this study, data from 996 patients with GC after gastrectomy were utilized. The data were split into groups, trained and tested, and a ratio of 8:2 was used to develop different machine learning models. Furthermore, the six different machine learning algorithms used in predicting the stage of GC include decision tree (DT), K nearest neighbor (KNN), logistic regression (LR), naive Bayes (NB), random forest (RF), and support vector machine (SVM) methods. Results: The analysis of the demographic variables revealed statistically significant differences in the PLR and NLR and other parameters between the two groups of patients with stages I and III gastric cancer (P < 0.05).

**Results:** The analysis of demographic variables revealed statistically significant differences in the PLR, NLR, and other variables between the two groups of patients with stages I and III gastric cancer, with a significance level of P-value < 0.05. Moreover, these findings suggest that the KNN model in this study is one of the best models for predicting the stage of GC.

## Introduction

Gastric cancer (GC) is the fourth most common malignancy and the second leading cause of death worldwide. Early gastric cancer has a good prognosis; however, it presents numerous unusual and unclear clinical symptoms (1, 2). More than 1 million cases of early gastric cancer are diagnosed each year. Early gastric cancer can be treated with endoscopy, whereas the only option for advanced gastric cancer is surgery (3). GC is usually asymptomatic in its early stages, and diagnosing GC during these stages is a challenging process. Nevertheless, early identification of patients is effective in improving their prognosis. Therefore, rapid, accurate, and reliable prediction is useful in optimizing the treatment of patients with GC (4). The 5-year survival rate for patients with GC is 90%, while this survival rate decreases to 18% for patients with stage III GC. Although the survival rate is high for surgeries performed in the early stages of the disease, patients with GC globally have an overall survival rate of 25% because most patients are diagnosed at advanced stages(3).

Predictive medicine is a rapidly evolving field that focuses on anticipating the onset of diseases before symptoms appear, enabling earlier and more personalized healthcare interventions (5). By leveraging data from genetics, lifestyle, environment, and advanced technologies such as machine learning and artificial intelligence, predictive medicine aims to identify individuals at high risk for specific conditions (6-8). This proactive approach allows healthcare providers to tailor prevention strategies, monitor high-risk patients more closely, and optimize treatment plans, ultimately improving patient outcomes and reducing healthcare costs (9-11). As the availability of large-scale health data continues to grow, predictive medicine holds significant promise for transforming the future of healthcare from reactive to preventative (12-14).

The adoption of artificial intelligence (AI)-based solutions, such as machine learning (ML), can overcome the limitations of invasive diagnostic procedures in the screening and diagnosis of GC due to their computational capabilities (15). ML-based techniques for developing predictive models and analyzing data rely on existing data and provide valuable insights from raw datasets (1, 15). ML models can make predictions on the basis of various measurements (16). ML methods have been applied with high accuracy to predict several types of cancer, including breast cancer and colorectal cancer (17).

Therefore, the use of artificial intelligence can optimally address intractable medical problems that were previously thought to be unsolvable. Identifying patients in the early stages of GC via highly accurate machine learning techniques is an efficient, rapid, and low-cost method that can assist physicians in diagnosing the disease stage more accurately and quickly (18, 19). The present study employed machine learning to predict various stages of GC in patients after surgery (20-22).

## Methods and Materials

### Study Design and Population

The present study included data from 1,331 patients with GC after gastrectomy, some of whom were excluded because of missing information. Therefore, this study was conducted on 996 patients. All patients were diagnosed with stage I-III GC on the basis of histological tests after surgery. Tumors were staged according to the TNM staging system of the 7th edition of the American Joint Cancer Commission (AJCC) (23). In this study, the inclusion and exclusion criteria included the absence of radiation or chemotherapy in the patients, the presence of complete clinical pathology and follow-up data of potential prognostic factors, the lack of other malignant cancers coexisting with the disease, the absence of GC recurrence, the absence of residual GC, and the lack of any acute inflammation or infection two weeks before surgery.

Data, including sex, age, tumor characteristics after surgery, necessary laboratory tests before surgery, and survival time, were then collected. Blood samples were taken from patients one week before surgery. The samples were divided into two groups: highly differentiated (papillary and moderately differentiated GC) and poorly differentiated (mucinous, signet-ring cell, and undifferentiated GC) (24). To monitor the patients, various examinations, including physical examinations, dynamic CT examinations, complete blood counts, gastroscopy, and measurements of tumor markers in the blood serum, were performed every three months for the first two years and every six months thereafter. The date of the last follow-up was considered June 2015 or the date of death due to any cause (20-22). Overall survival (OS) was defined as the time interval from the date of surgery to the date of the patient’s last follow-up or death from any cause.

### Data availability

Data from this study are available in the BioStudies database: https://www.ebi.ac.uk/biostudies/studies?query=S-EPMC5373584

### Calculations for Biomarkers

The NLR is obtained by dividing the absolute number of neutrophils by the absolute number of lymphocytes, and the PLR is obtained by dividing the absolute number of platelets by the absolute number of lymphocytes (25). COP-NLR was calculated on the basis of previous studies, with a score of 0 or 1 assigned to patients with normal values and a score of 2 assigned to patients with high platelet counts (> 300 × 10^9 /L) and a neutrophil□lymphocyte ratio greater than 3 (26). Finally, the prediction of the disease stage will be made based on the data available in two phases, including the initial phase of GC, stage I, and the final phase of GC, stage III.

### Machine Learning Algorithms

This study used six ML algorithms to predict the stage of GC, including DT, KNN, LR, NB, RF, and SVM (27-30). The DT algorithm is a model of classical supervised learning that uses a tree diagram structured like a flowchart (31). The KNN algorithm is a nonparametric, supervised learning model that utilizes the nearest neighboring individuals in the population for identification (32). LR employs a logistic function to model a dependent binary variable and is also a tool for statistical measurements and regression analyses (33). NB is a statistical technique that applies Bayes’ theorem for classification (34). It is one of the simplest ML algorithms, examine a feature in a class independently of other features. RF is an algorithm that uses the results of multiple DTs to achieve a specific output (35). SVM is a supervised ML algorithm that classifies data via an optimal line or plane (36).

### Machine Learning Data Processing Methods

First, the data were analyzed on the basis of descriptive statistics. A P-value <0.05 was considered significant. The necessary analyses were performed to investigate different ML algorithms via the R programming language (V.4.3.1). For this purpose, 80% of the data were used for training, and the other 20% were used for testing to investigate the prediction of the GC stage.

## Results

### Demographic characteristics of the study population

The study population included 1,331 patients with GC, of whom 996 patients were used to design the model: 220 patients in stage I (65 women and 155 men) and 776 patients in stage III (258 women and 518 men). The remaining studies were excluded from the analysis because of a lack of information. The mean age of the study population with stage I GC was 56.10 ± 12.60 years, and for stage III GC, it was 57.77 ± 11.50 years (Table 1).

**Table 1:**
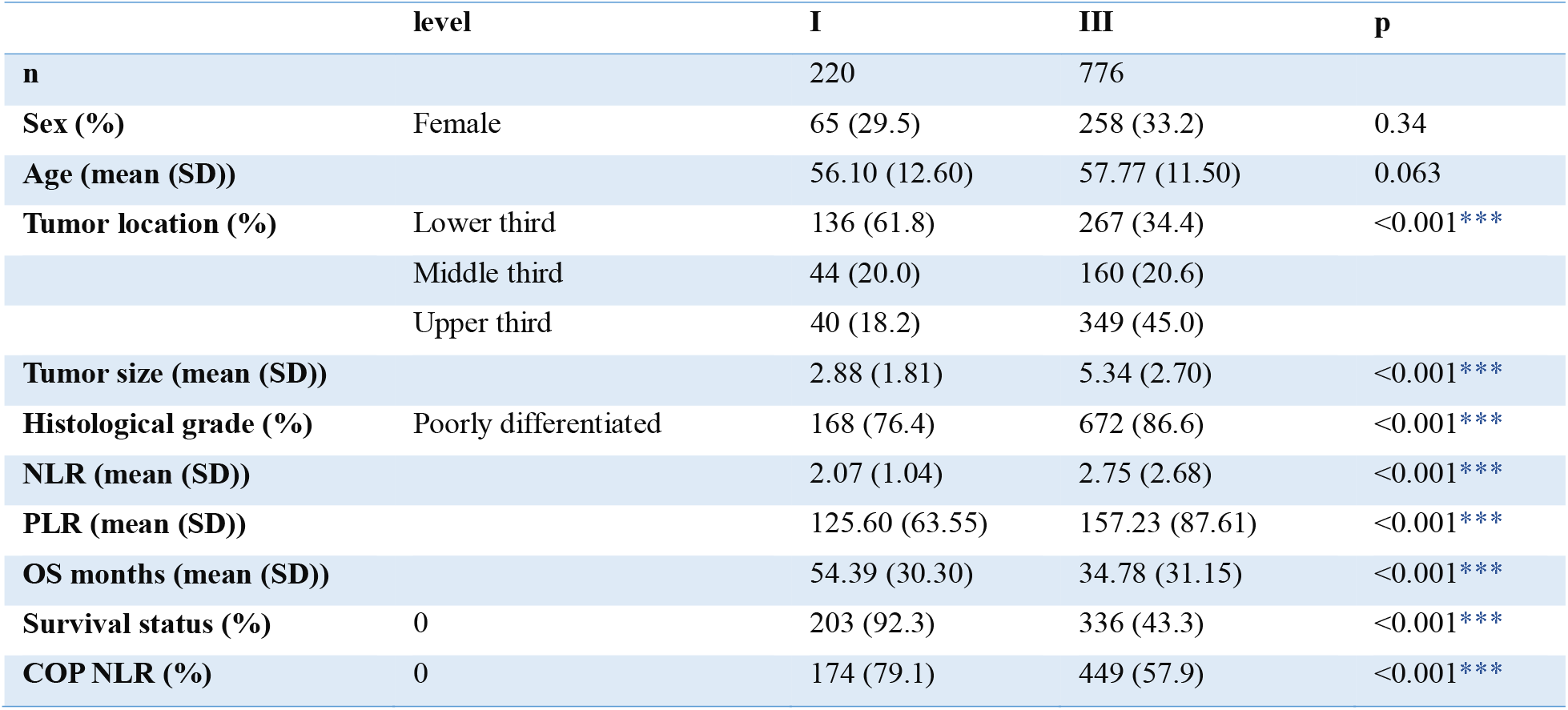
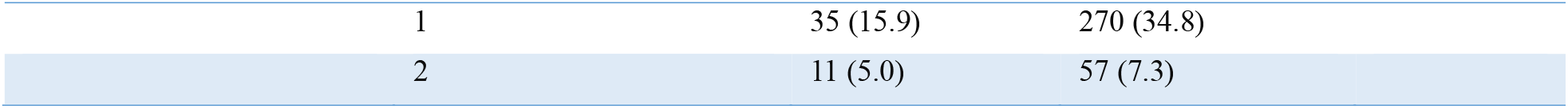
Basic data of variables and their significance levels between two different stages of GC

The results of the comparison of the two groups in stages I and III of GC revealed that age and sex were not significant demographic criteria in the disease stage. However, the PLR, NLR, and other characteristics were significantly different between the two groups in stages I and III of GC (P < 0.05) (Table 1). Patients with stage III GC exhibit more advanced disease characteristics, such as larger tumor sizes, higher NLRs and PLRs, and poorer histological differentiation. Additionally, these patients had worse overall survival and higher mortality rates than patients with stage I GC did (Table 1).

### Analyzing Machine Learning

The results in Figure 2 show the performance of the six ML classifier algorithms on the basis of the selected features. The results in Figure 2 indicate that the performance of the KNN classifier is better than that of the other five algorithms. The detection accuracy of the SVM algorithm was excellent, with a high sensitivity of 0.97. This algorithm works effectively for all stages of cancer and results in few false negatives. However, since its specificity is 0.48, it misclassifies healthy cases.

**Figure 1.**
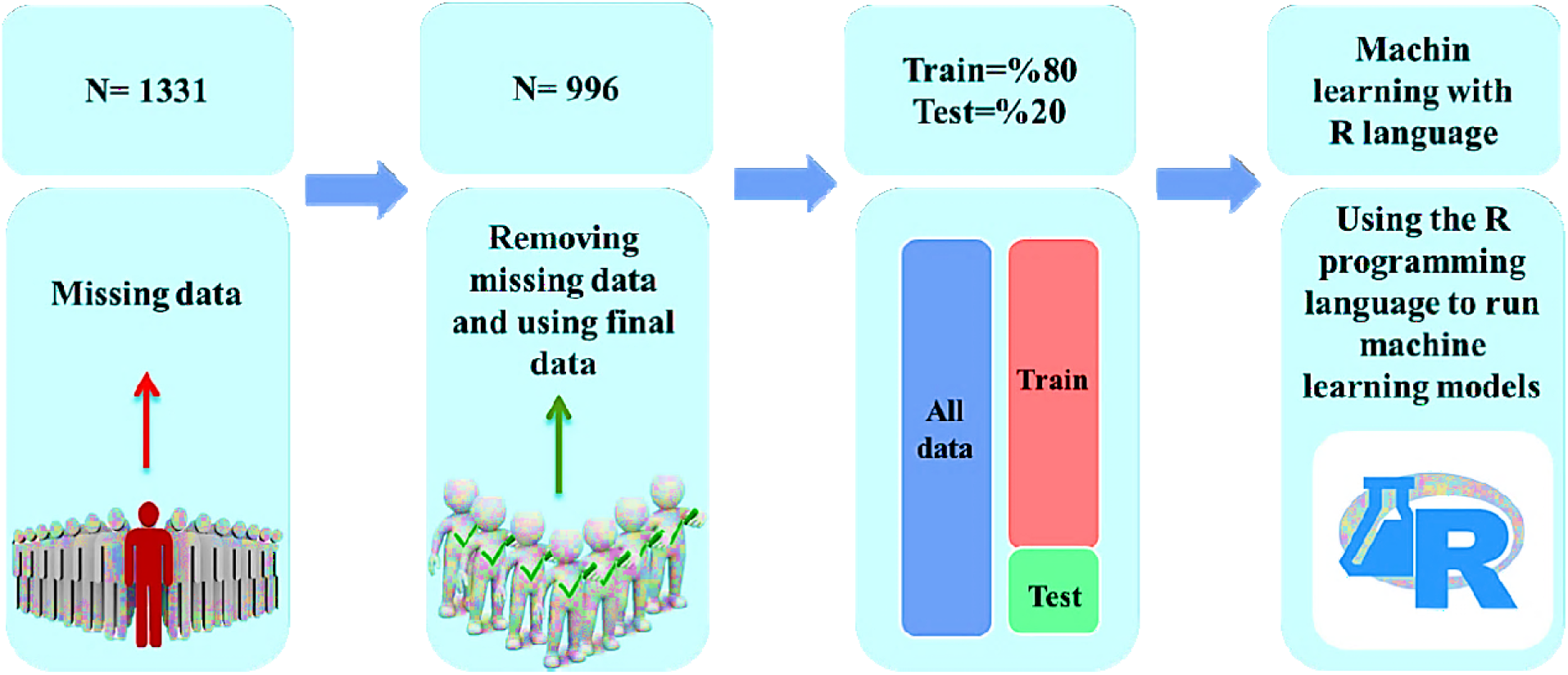
Overview of the steps involved in developing a GC staging prediction model

**Figure 2.**
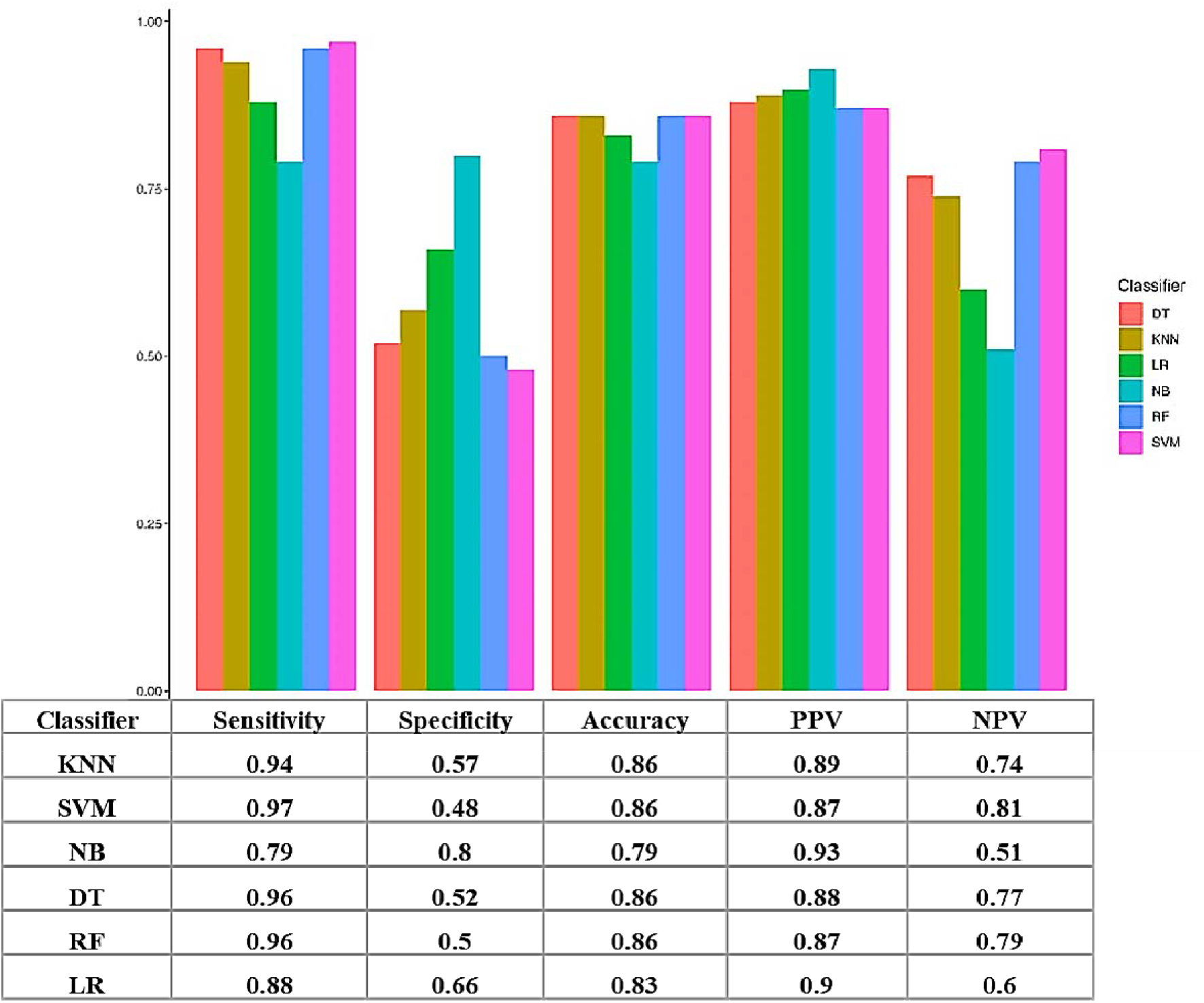
Rectangular diagram of six ML algorithms, including DT, KNN, LR, NB, RF, and SVM, along with their performance comparison.

NB is another algorithm that is suitable in terms of specificity (0.80) and positive predictive value (PPV = 0.93); it is used to confirm true positives and is reliable. However, because of its sensitivity (0.79), it is not effective in detecting actual cases of cancer. Although KNN, DT, and RF achieved balanced performances, with an above-average accuracy of 0.86 and sensitivity, they presented relatively low to moderate specificity. In the trade-off of specificity to sensitivity, SVM stands the best bet; however, to reduce the incidence of false positives, the RF or DT models would be better candidates since they delivered balanced performance (Figure 2).

### Feature Analysis

This analysis studied the feature contribution to the RF model for predicting the GC stage. The importance of a feature is generally expressed by the %IncMSE-rank: the higher, the more important a feature is when the model performance decreases significantly due to changes in the feature values. The top ten percent features show, in order of importance, the most important features contributing to GC. Tumor size and survival status appeared to be the most important features. The importance of features decreased from top to bottom (Figure 3).

**Figure 3.**
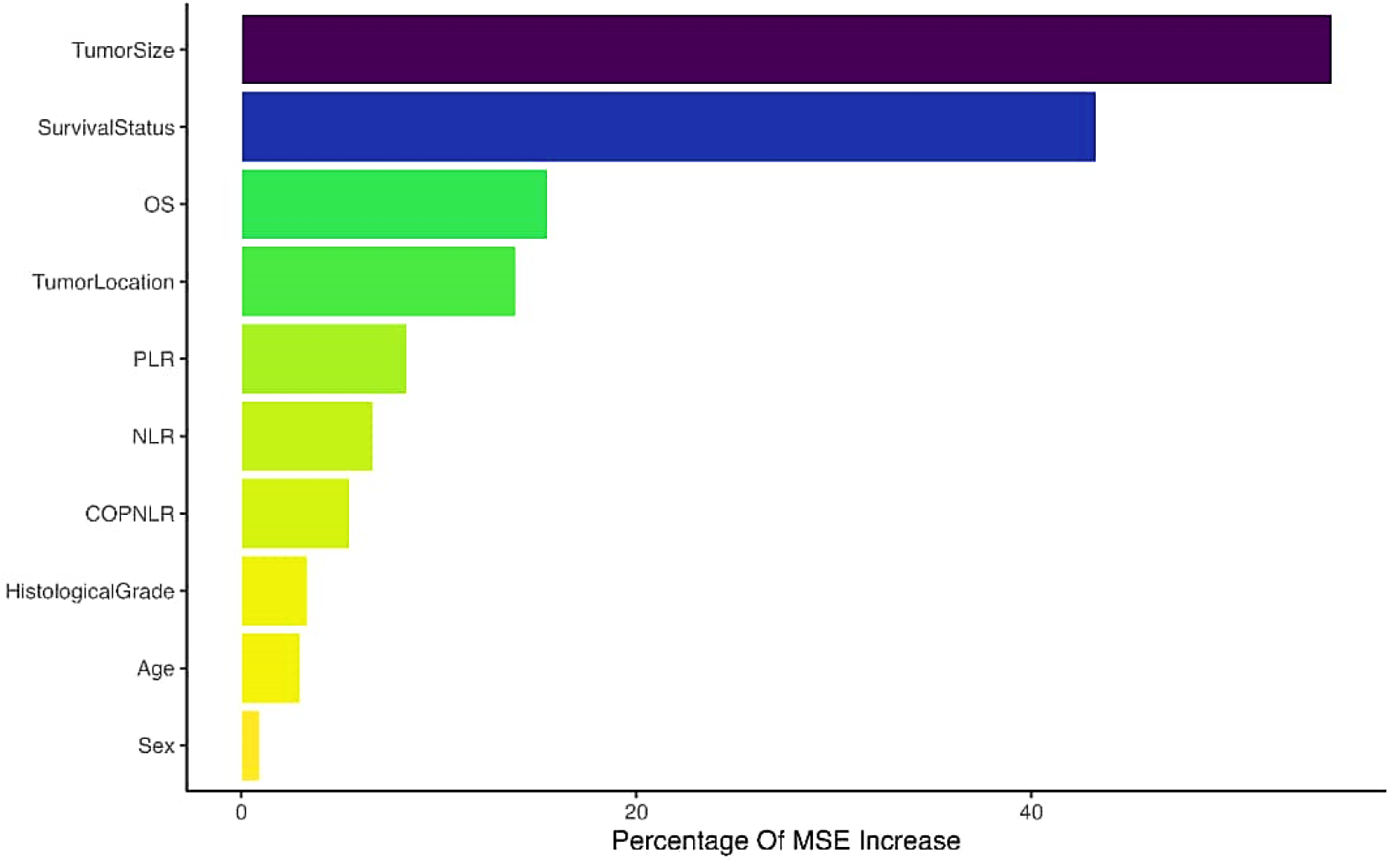
Important features for predicting disease stage in patients with GC after surgery, IncMSE (percentage increase in the mean squared error)

## Discussion

In this study, based on the available data for diagnosing stage I and stage III GC, six different ML algorithms were used to predict the stage of GC. Compared with other ML algorithms, the KNN algorithm provided appropriate results and reported more balanced outcomes. Additionally, the stage of GC had a statistically significant relationship with various variables in this study, including tumor location, tumor size, histological grade of tissue, NLR, PLR, OS months, survival status, and COP NLR. Among these variables, tumor size and survival status were the most important features associated with disease stage. However, just as sex and age did not have a statistically significant relationship with cancer stage, they were deemed the least important features.

GC is one of the most common and deadly tumors globally, primarily because patients with GC are often diagnosed at advanced stages of the disease. Therefore, efforts to develop noninvasive screening methods are necessary to increase patient survival and quality of life (37-39). Studies indicate that several factors affect the disease stage and prognosis of patients with GC, including tumor size and tumor location (40). The results of this study also emphasize this point, as tumor location and tumor size are significantly correlated with disease stage. Furthermore, as shown in Figure 3, the results indicated that the most important feature was tumor size.

Several biomarkers of the inflammatory response in cancer, including the neutrophil-to-lymphocyte ratio (NLR) and the platelet-to-lymphocyte ratio (PLR), which are crucial prognostic indicators, are important prognostic factors (41). PLR is a particularly significant factor in stage III/IV CRC. Compared with those of tumor markers, the levels of systemic inflammatory markers are notably increased in stage I GC patients. Consequently, inflammatory markers present in peripheral blood are vital for the early diagnosis and screening of GC (42). These data confirmed the results of the present study, demonstrating that biomarkers such as the PLR and NLR are significantly related to the stage of GC. However, one study indicated that NLR and PLR markers were more strongly associated with the diagnosis of GC in male patients, suggesting that sex plays a role in this context (43). In contrast, the present study showed no relationship between sex or biomarkers and the stage of the disease.

L. Lian et al. investigated the NLR and PLR and their relationships with GC prognosis and reported that the NLR and PLR in patients with GC were significantly greater before surgery than they were in healthy subjects. Their results revealed that lower levels of two factors, PLR and NLR, before surgery indicated less damage, including a lower depth of invasion and fewer lymph node metastases (44). Zhou et al. demonstrated that GBM was the best model for predicting the prognosis of inflammatory factors in GC; however, the model still requires further training to achieve greater accuracy (23). In this study, like our study, models such as LR were also utilized, with the accuracy of the current study being slightly higher in this model. Xu et al. also predicted colorectal cancer recurrence via DT, gradient boosting, LR, and GBM algorithms, among which the well-developed algorithms were gradient boosting and GBM (42). The current study did not use the GBM algorithm but instead employed other algorithms. In a study aimed at predicting bladder cancer stage identification via ML, Kouznetsova et al. reported that MLP_PCA_1 has a high average accuracy (45).

In summary, the present study investigated the relationships between various features and the stage of GC and demonstrated that tumor size, survival status, overall survival status, and tumor location are the most important effective features in the disease stage. Additionally, among the algorithms used, the KNN algorithm is one of the best, exhibiting the highest accuracy, along with the SVM, DT, and RF algorithms, while it is more specific than the other algorithms. Therefore, the use of ML is a noninvasive and efficient method for predicting the stage of GC.

## Data Availability

All data produced are available online at

https://www.ebi.ac.uk/biostudies/studies?query=S-EPMC5373584

## Abbreviations

GC: Gastric cancer
ML: Machine Learning
DT: Decision Tree
KNN: K Nearest Neighbor
LR: Logistic Regression
NB: Naive Bayes
RF: Random forest
SVM: Support Vector Machine

